# Photoacoustic imaging of intestinal physiological biomarkers in infants and children

**DOI:** 10.64898/2026.09.14.26362277

**Authors:** Paulina Wright, London Paige, Sandra Norona, Cherrie D. Welch, Kristen A. Zeller, Victoria G. Weis, Jared A. Weis

## Abstract

**Background:** A major hallmark of necrotizing enterocolitis (NEC) is intestinal hypoxia, but detection of this feature is difficult. Photoacoustic imaging (PAI) is an emerging imaging method to noninvasively visualize intestinal oxygenation and microvasculature and is promising for improved NEC detection. Prior to clinical translation of PAI for NEC detection, initial studies must be performed in a pediatric population to assess safety and feasibility.

**Methods:** Under IRB approval, eighteen healthy pediatric participants underwent PAI scans before and after eating a lipid-rich snack. PAI was acquired in each abdominal quadrant with intestinal regions-of-interest (ROI) manually drawn on each image series to measure averaged values of deoxygenated hemoglobin (Hb), oxygenated hemoglobin (HbO_2_), total hemoglobin (HbT), and oxygen saturation (sO_2_).

**Results:** Minor temporary erythema and pressure from safety goggles in some participants were the only adverse events reported. Pre-/post-prandial changes for HbT, HbO2, Hb, and sO2 were assessed for each abdominal region and age cohort. Results showed significant post-prandial hyperemia in the intestines with no changes in bowel oxygenation.

**Conclusions:** PAI is a safe and feasible imaging modality for evaluating intestinal blood flow in pediatric patients that detects physiological changes in bowel microvasculature and shows strong potential as a quantitative biomarker for assessment and monitoring of intestinal vascular health.

**Highlights:**

- Pediatric abdominal PAI is safe: Only minor, temporary, and immediately reversible adverse events were reported due to goggles
- Pediatric abdominal PAI is feasible: Excellent sensitivity for detecting changes in intestinal microvascular biomarkers
- First-in-early-pediatric study: Our findings support the translational use of PAI for assessing intestinal vascular health

## Introduction

Necrotizing enterocolitis (NEC) is a rare but devastating neonatal disease that primarily impacts very low birthweight (VLBW) infants within the first few weeks of life [1, 2]. Trends for infants show that lower birth weight and younger gestational ages are more likely to be diagnosed with NEC, require surgery, and have higher mortality rates and worse long-term outcomes [1–6]. Some hallmarks of NEC include pneumatosis intestinalis, pneumoperitoneum, hypoxia, reduction in peristalsis, and ischemia (5-8).

Current diagnostic strategies begin with monitoring for clinical signs and symptoms of feeding intolerance, bloody stool, abdominal distention, and vomiting [6–8]. Upon suspicion, serial abdominal plain-film radiography (AXR) is used to detect changes in visualized GI anatomy [3, 9–12]. These methods lack diagnostic accuracy due to low sensitivity and specificity [7, 8, 11]. While bowel ultrasound (BUS) is emerging to improve diagnostic specificity, sensitivity remains lacking, as BUS is often reliant on observer experience with NEC, which can cause inequity of care [7–15]. Near-infrared spectroscopy (NIRS) has also been explored to improve NEC detection and diagnosis [8, 12, 16–21]. NIRS measures tissue oxygenation as a single-point measurement and has shown some mixed success in both animal models and clinical studies but is limited by lack of tissue penetration [16–18, 21–23]. NIRS also yields single-point measures and does not allow for spatial visualization of tissue oxygenation, limiting interpretability of the measurement with potential error due to proximity to large blood vessels [12, 16, 17, 20, 21].

Photoacoustic imaging (PAI) is an emerging imaging modality for which we have previously shown success in detecting hallmarks of NEC in preclinical animal studies [24–29]. PAI uses a unique ‘light-in and sound-out’ method to visualize and measure chromophores within tissues [17, 30–32]. With PAI, infrared light pulses at specified wavelengths penetrate tissue and excite chromophores within tissue that undergo thermoelastic expansion and acoustic signal production that is detected by an ultrasound probe [30–33]. Using computational spectral unmixing, concentrations of specific chromophores can then be determined, including oxyhemoglobin and deoxyhemoglobin [31, 34, 35]. Previously, we demonstrated in a neonatal rat pup animal model that PAI is sensitive to changes in intestinal tissue oxygenation [26]. In a subsequent neonatal rat pup model of experimental NEC, we demonstrated that PAI can detect differences in intestinal tissue oxygenation between NEC and breastfed control animals [24, 25, 27]. In a recent clinical study, Regensburger et al. demonstrated PAI in pediatric abdominal tissue to assess inflammatory bowel disease activity, with pediatric subjects of various ages down to 3-years [33]. Combined, these studies show significant promise for the use of PAI to improve detection of NEC as PAI has been shown to be sensitive to changes in intestinal microvasculature and hypoxia, major hallmarks of NEC [24–27, 33]. However, there is a gap in our current knowledge of safety and feasibility for intestinal PAI in younger aged participants. As we work towards clinical translation for NEC diagnosis, there is a need to address these gaps in younger pediatric participants.

In this study, our goal is to test the safety and feasibility of abdominal PAI in a cohort of healthy pediatric participants. We conducted paired pre- and post-prandial PAI imaging sessions to determine the sensitivity of PAI to the known physiological hyperemic response from digestion of a lipid-rich snack [20, 22, 36–40]. This study is an important translational step forward towards our ultimate goal for the use of PAI within the NICU population as a biomarker of intestinal vascular health and improved NEC diagnosis.

## Methods

### Participants and Experimental Design

This prospective pilot study was performed under Wake Forest University School of Medicine IRB approval. A schematic of the study design is shown in Figure 1. Two-parent/guardian consent was obtained for all participants, and assent was obtained for children aged 7 and older. Age, height, weight, gender, and race were recorded for each participant. Participants were excluded if they had participated in an interventional study six months prior, had a current diagnosis or history of cancer, autoimmune disease, metabolic or nutritional disorders, GI surgery, or illnesses or disorders including but not limited to celiac disease, Crohn’s disease, Hirschsprung’s disease, gut motility disorders, and IBD. PAI scans were performed with participants in a supine or reclined seated position both before and after eating a lipid-rich snack (ice-cream, breast milk, or formula based on age and parental preference) with 30 - 60 minutes between scans. Scans were acquired in each of the four abdominal quadrants, when possible, due to patient size: Left Upper Quadrant (LUQ), Right Upper Quadrant (RUQ), Right Lower Quadrant (RLQ), and Left Lower Quadrant (LLQ). For younger/smaller participants, only two abdominal regions could be acquired due to the size of the imaging probe relative to the abdomen: Upper and Lower. For each participant, ultrasound gel was used to provide acoustic coupling with the abdomen of participants. Participants were provided with safety goggles for eye protection from the laser used in PAI imaging, and the study team verified sizing and adjusted for fit as needed. Participants were given a safety assessment questionnaire after each PAI imaging session to determine participant comfort, visual assessment of tissue changes, and other contraindications.

**Figure 1.**
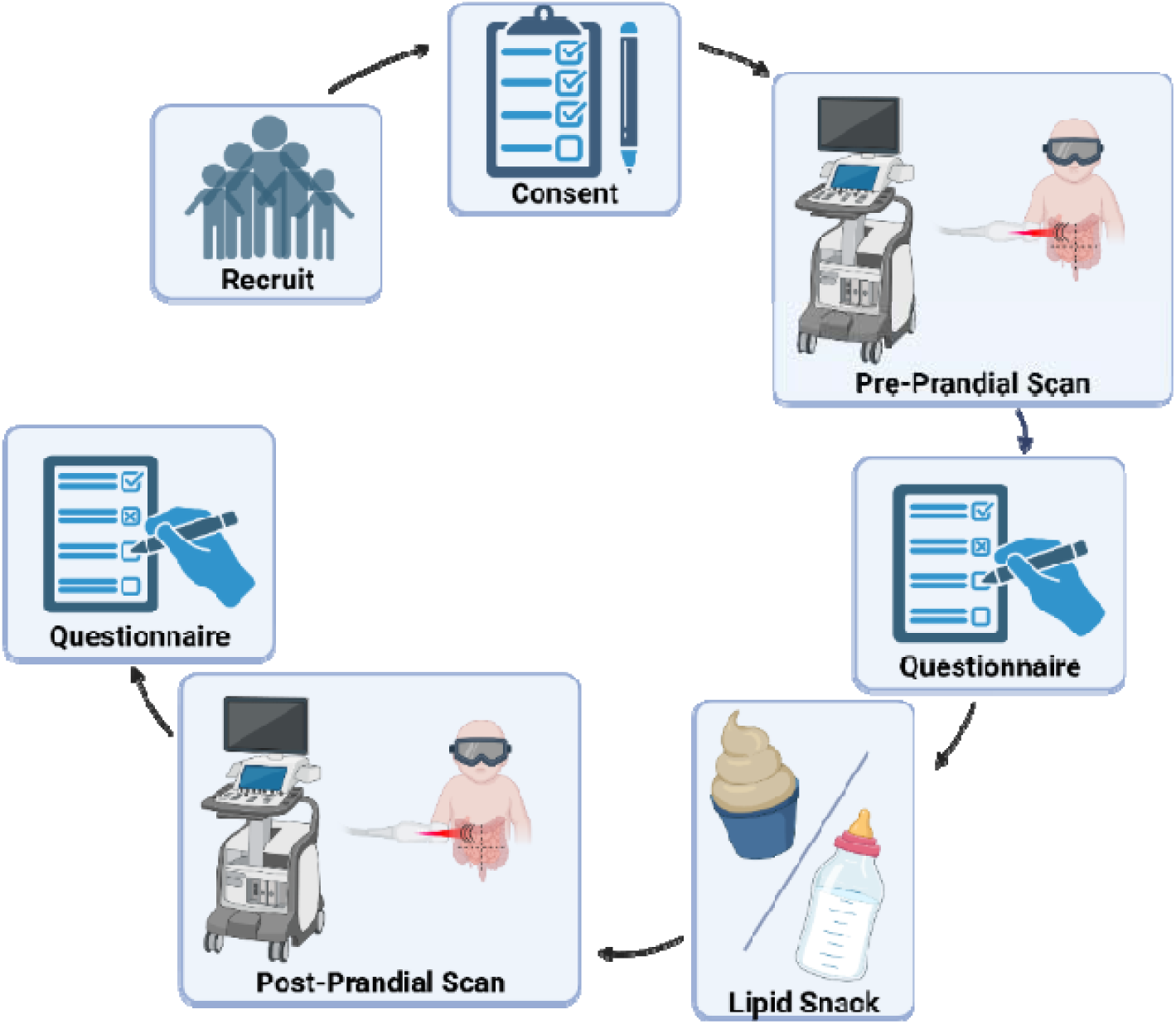
Schematic representation of study design, showing the process for participant recruiting, consenting, participating in the pre-prandial scan, answering the first safety questionnaire, eating the lipid-rich snack, then waiting thirty minutes to an hour for digestion, participating in the post-prandial scan, and answering the second safety questionnaire.

### Ultrasound and Photoacoustic Imaging

An iThera MSOT Acuity (iThera Medical, Munich, Germany) system was used for imaging using a pre-defined ‘Oxygenation’ preset that utilizes 7 wavelengths: 700, 730, 760, 780, 800, 850, and 900 nm. The iThera system is coupled with ultrasound to provide 2D B-mode ultrasound images to visualize anatomical structures. Each abdominal quadrant was imaged with cine video recordings of approximately 30 seconds.

### Reconstruction and Spectral Unmixing

Images were reconstructed using the ViewMSOT software (version 4.0.3.4, iThera Medical) with a back projection algorithm. Fluence correction using default settings was used to compensate for depth-dependent decay of optical fluence [34]. Assessment of intestinal tissue total hemoglobin (HbT), oxyhemoglobin (HbO_2_), deoxyhemoglobin (Hb), and oxygen saturation (sO_2_) is based on automatic spectral unmixing calculation of oxy- and deoxyhemoglobin concentrations within ViewMSOT postprocessing analysis software based on known spectral differences of Hb and HbO_2_. HbT is calculated as the sum of Hb and HbO_2,_ and sO_2_ is calculated as the ratio of HbO_2_ and HbT. Mean HbT, HbO_2_, Hb, and sO_2_ measurements were quantified within manually designated regions-of-interest (ROI) drawn over the intestinal region at depths with adequate PAI signal intensity based on the anatomical ultrasound images. Tissue depth was limited by poor signal intensity beyond approximately 2 cm for photoacoustic measurements.

### Adverse Event Assessment Questionnaire

After each pre-prandial and post-prandial imaging session, participants were administered a questionnaire to assess adverse events (AE). Participants were asked questions to assess the presence or absence of any pressure, pain, irritation, or redness from the laser safety glasses. Participants were also asked to assess the presence or absence of any pressure, pain, skin irritation, redness, tingling, numbness, coolness, or heat from the imaging probe. Safety assessment information was collected after each imaging session using participant questionnaires when feasible; in infants and younger participants, data were obtained from parent or surrogate interviews and imaging technician observations. A visual assessment of both the abdominal and periorbital regions was also documented by the study team. All reported AE were recorded for when the symptoms occurred and when symptoms concluded. If the event did not conclude during the session, follow-up contact was made to track resolution of the event.

### Statistical Analysis

Primary analysis included all participants enrolled with evaluable images that contained PAI signal intensity at intestinal tissue depth. Pre-to-post-prandial changes in mean HbT, HbO_2_, Hb, and sO_2_ measurements were analyzed using paired two-tailed t-tests both within each region and averaged across all regions. For participants under 1-year, the relative size of the imaging probe limited our ability to scan within abdominal quadrants; therefore, upper and lower abdominal regions were substituted, with measures in upper/lower regions repeated in analysis for consistency with quadrant definitions. Metrics within the upper region were repeated for both RUQ and LUQ, and metrics within the lower region were repeated for both RLQ and LLQ in these participants.

Secondary analysis examined pre-to-post-prandial changes in mean HbT, HbO_2_, Hb, and sO_2_ measurements within predefined age cohorts (0-2 years, 3-6 years, and 7-12 years) using paired two-tailed t-tests within each age cohort.

## Results

Eighteen healthy participants aged 6 weeks to 12 years old were enrolled, as in Table 1. Two participants were excluded from statistical analysis due to insufficient optical penetration depth in subjects greater than 10-years-old due to body habitus. The primary analysis included all sixteen participants with metrics analyzed within abdominal quadrant locations. In a secondary analysis, metrics were analyzed within age-based cohorts divided into three age groups: 0-2 years (n=6), 3-6 years (n=6), and 7-9 years (n=4).

**Table 1.**
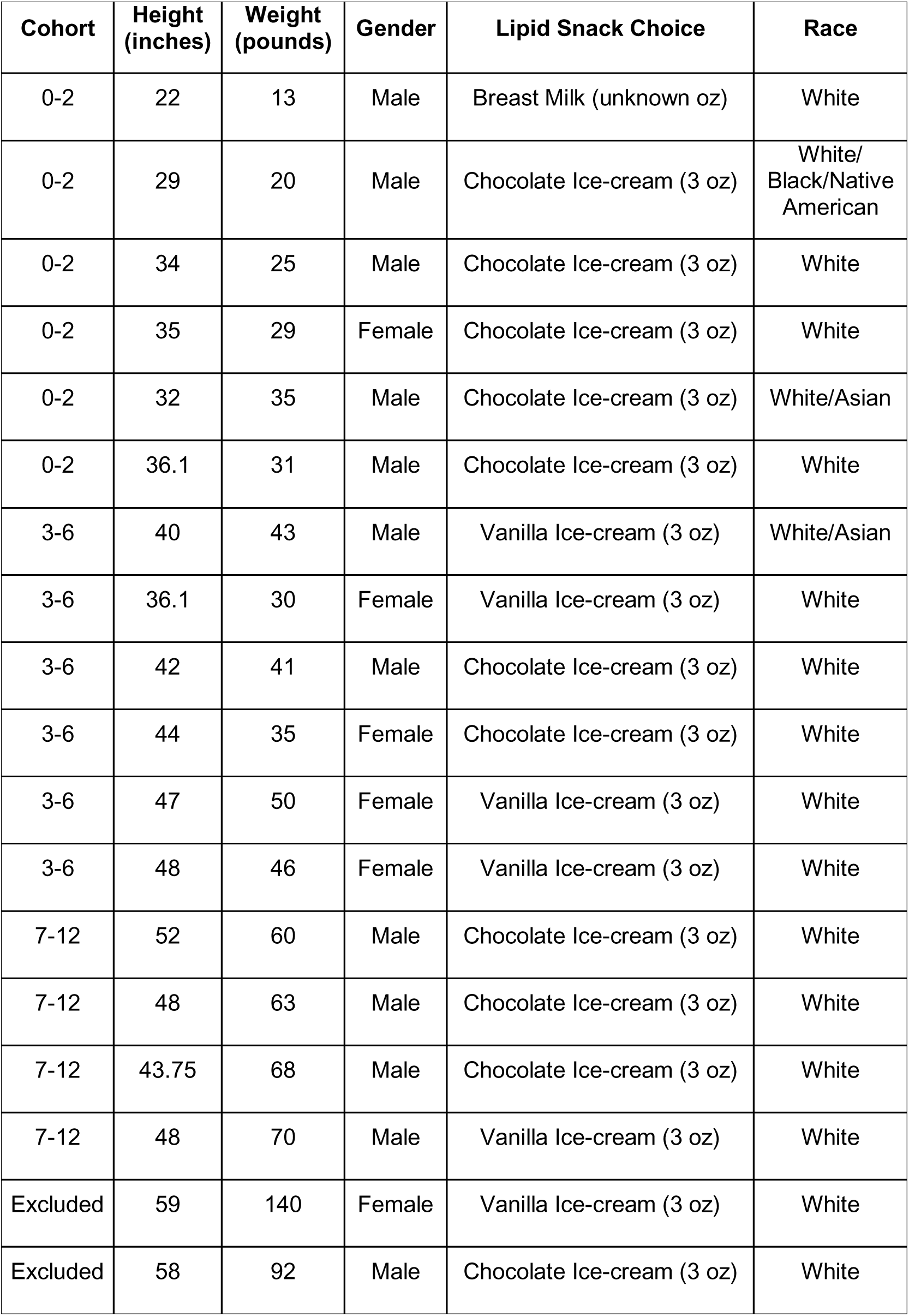
Demographic information of participants including cohort, height, weight, gender, snack choice, and race.

### Primary Analysis of Abdominal Quadrants Across the Entire Cohort

Representative images from pre- and post-prandial PAI imaging sessions for participants from each age cohort (0-2 years, 3-6 years, and 7-9 years) are shown in Figures 2-4. In Figure 2, PAI images from a participant in the 7-9-year cohort show a relative increase in HbT, HbO_2_, and Hb intensity from the pre- to post-prandial imaging session, with sO_2_ intensity maintained between the imaging sessions. In Figure 3, images from a participant in the 3-6-year cohort show a similar pattern of intensity changes with a slight increase in visualization depth in the younger/smaller participant. In Figure 4, images from a participant in the 0-2-year cohort show similar patterns of PAI metric intensity increase and demonstrate deeper visualization of intestinal tissues in the much smaller/younger participant. Scan averages were created by averaging all regional values for an individual participant in each imaging session. Scan averages were divided into cohorts based on regional location in the abdomen. Quantitative pre- and post-prandial measures within each abdominal quadrant for HbT, HbO_2_, Hb, and sO_2_ are shown in Figure 5 for all participants, with pre-/post-prandial changes shown as means of differences (µ) and standard deviation of differences (std). Quantitative pre- and post-prandial measures for the averaged intestinal regions for HbT, HbO_2_, Hb, and sO_2_ are shown in Figure 6. Overall, significant increases are observed from the pre- to post-prandial imaging session for HbT, HbO_2_, and Hb, whereas sO_2_ changes are insignificant. Specifically, significant differences between the pre- and post-prandial imaging session were found for HbT in the LUQ (µ = 0.3240, std ± 0.4685, p < 0.05), RUQ (µ = 0.2588, std ± 0.3286, p < 0.01), RLQ (µ = 0.2308, std ± 0.3203, p < 0.05), and averaged regions (µ = 0.2452, std ± 0.2312, p < 0.001). Significant values were found for HbO_2_ in the LUQ (µ = 0.1981, std ± 0.3250, p < 0.05), RUQ (µ = 0.1761, std ± 0.2338, p < 0.01), RLQ (µ = 0.1547, std ± 0.2660, p < 0.05), and averaged regions (µ = 0.1601, std ± 0.1726, p < 0.01). Significant values were found for Hb in the LUQ (µ = 0.1259, std ± 0.1516, p < 0.01), RUQ (µ = 0.08264, std ± 0.1126, p < 0.05), RLQ (µ = 0.07604, std ± 0.08583, p < 0.01), LLQ (µ = 0.05597, std ± 0.07280, p < 0.01), and averaged regions (µ = 0.08513, std ± 0.07636, p < 0.001). No significant values were found for sO_2_ in any region.

**Figure 2.**
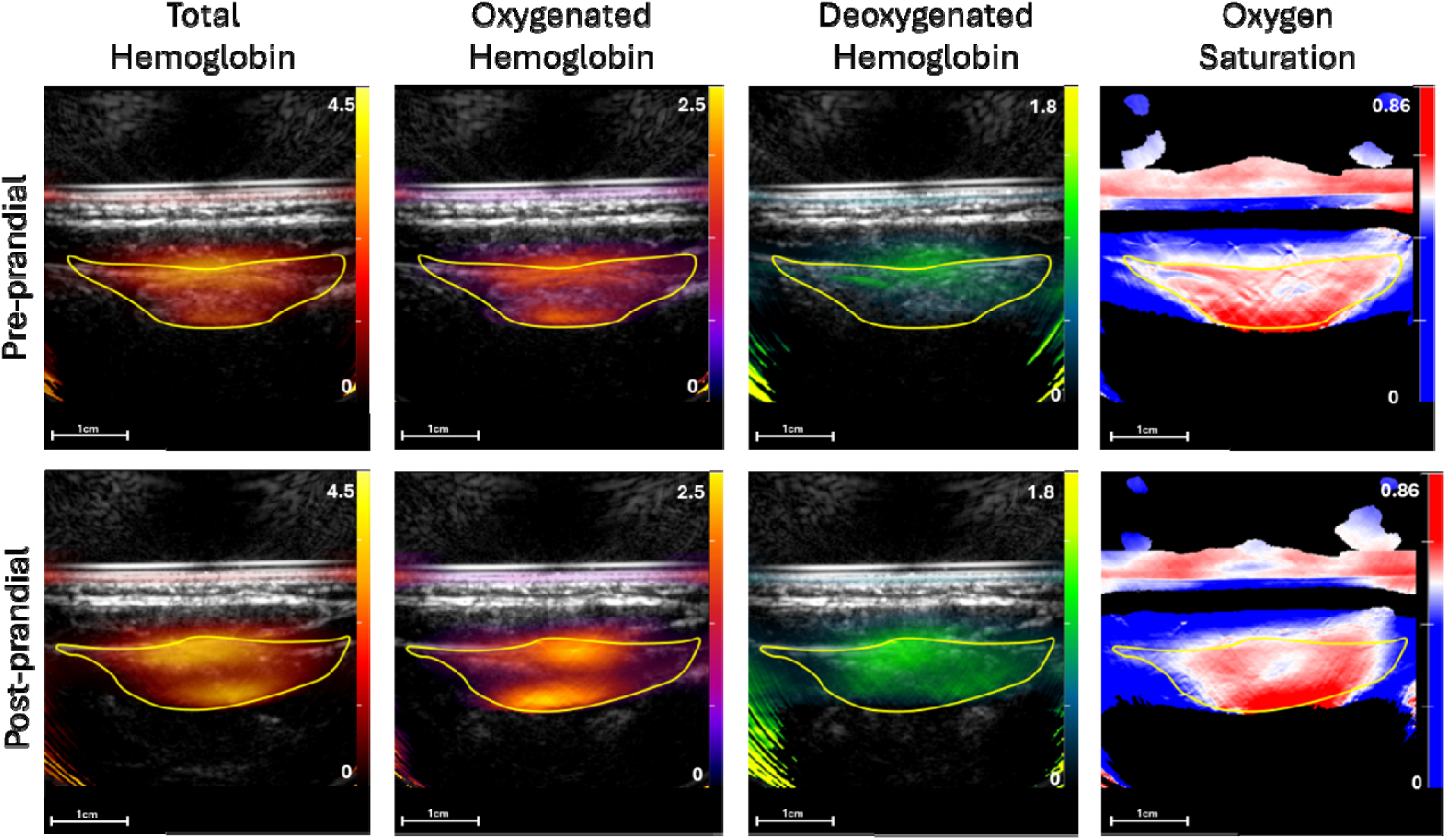
Representative photoacoustic images overlaid on ultrasound images of a participant from the 7-9 years-old cohort. Pre- and post-prandial images for total, oxy-, deoxyhemoglobin, and oxygen saturation are shown within the LUQ. Regions of intere**s**t from the central intestinal region are used for measurement. A post-prandial increase can be seen visually by the increase in brightness indicated by the color bar within the ROI for HbT, HbO2, & Hb. Scale bar = 1cm.

**Figure 3.**
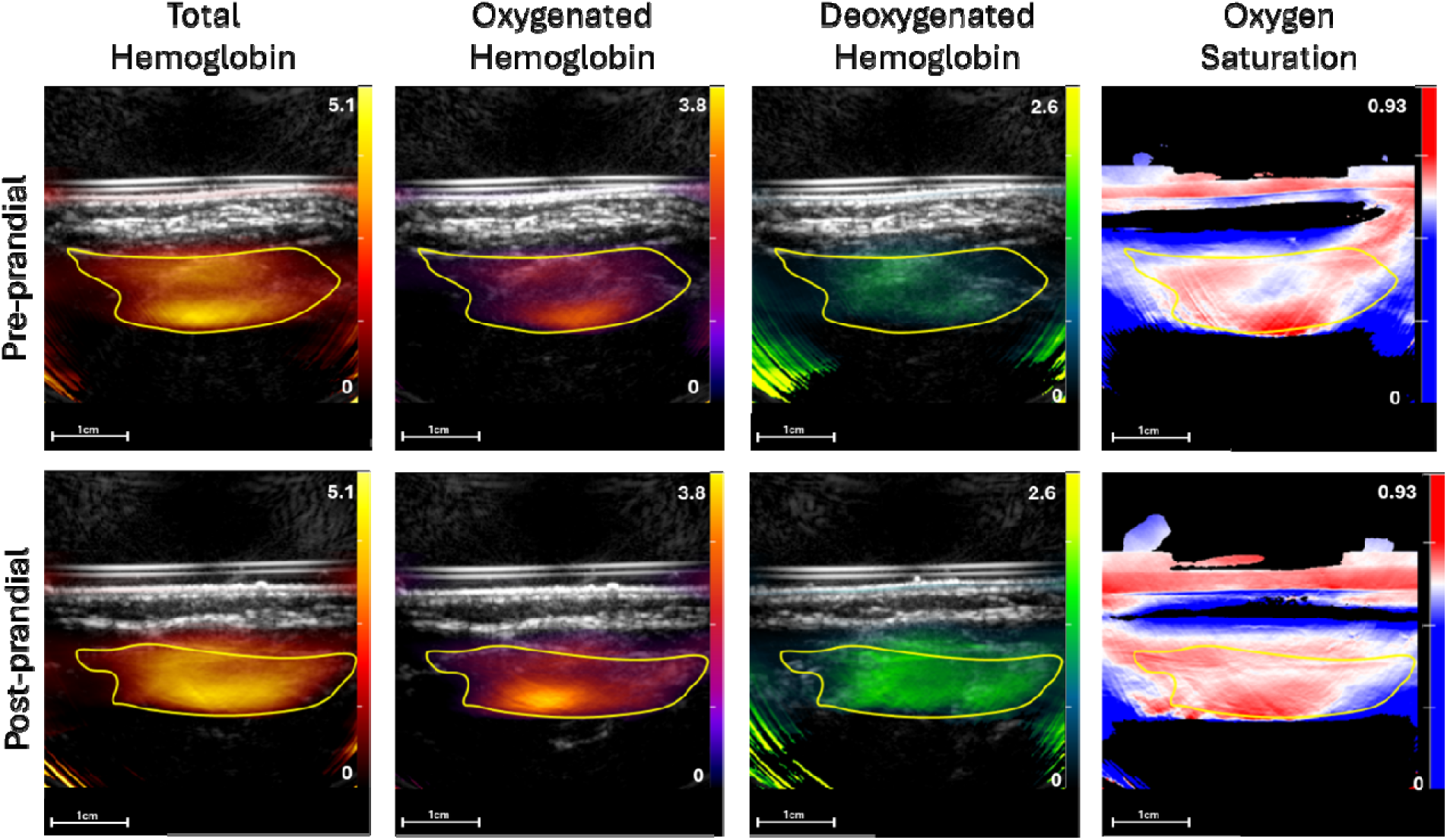
Representative photoacoustic images overlaid on ultrasound images of a participant from the 3-6 years-old cohort. Pre- and post-prandial images for total, oxy-, deoxyhemoglobin, and oxygen saturation are shown within the RLQ. Regions of intere**s**t from the central intestinal region are used for measurement. A post-prandial increase can be seen visually by the increase in brightness indicated by the color bar within the ROI for HbT, HbO2, & Hb. Scale bar = 1cm.

**Figure 4.**
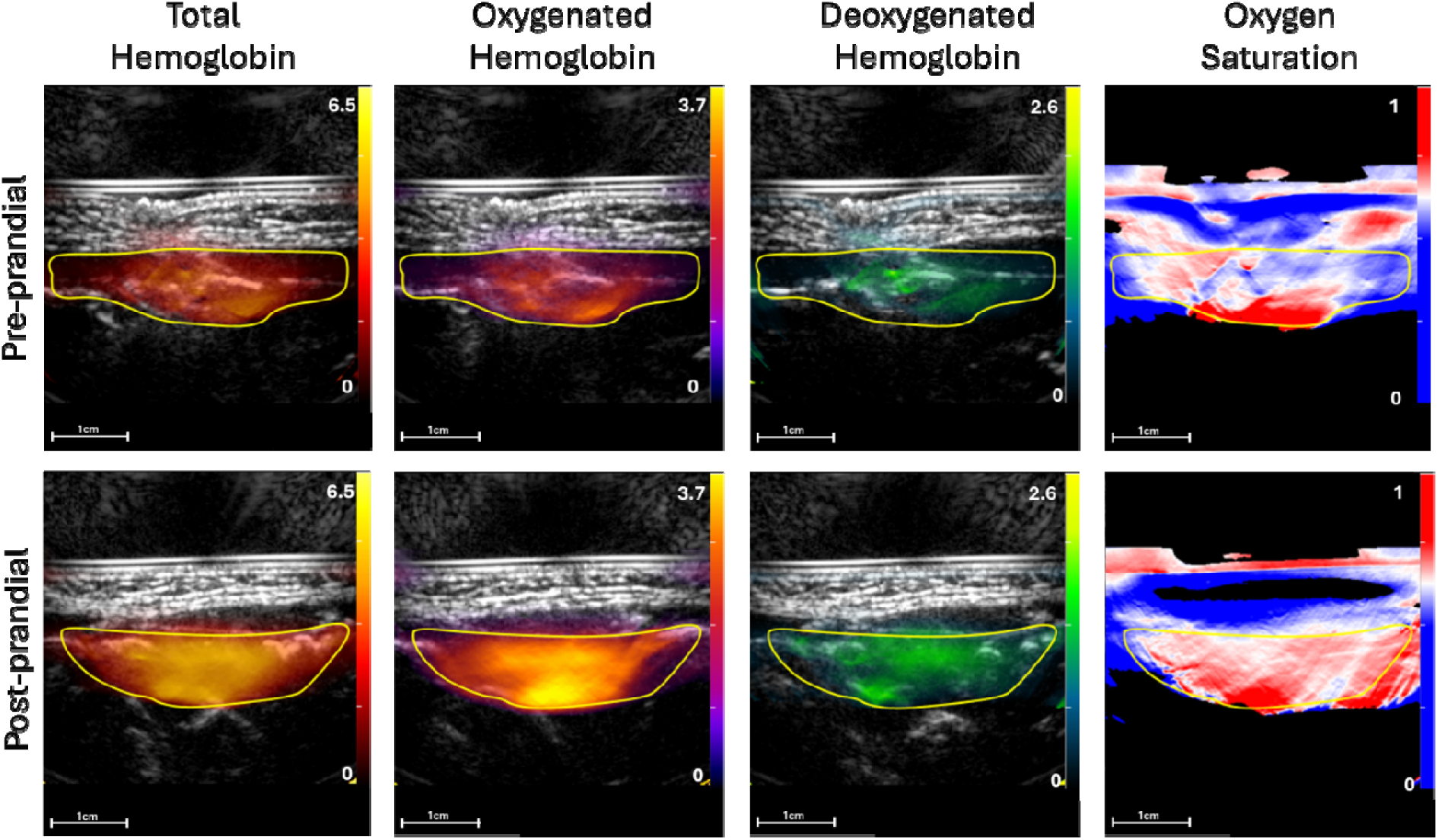
Representative photoacoustic images overlaid on ultrasound images of a participant from the 0-2 years-old cohort. Pre- and post-prandial images for total, oxy-, deoxyhemoglobin, and oxygen saturation are shown within the Lower Region. Regions of interest from the central intestinal region are used for measurement. A post-prandial increase can be seen visually by the increase in brightness indicated by the color bar within the ROI for HbT, HbO2, & Hb. Scale bar = 1cm.

**Figure 5.**
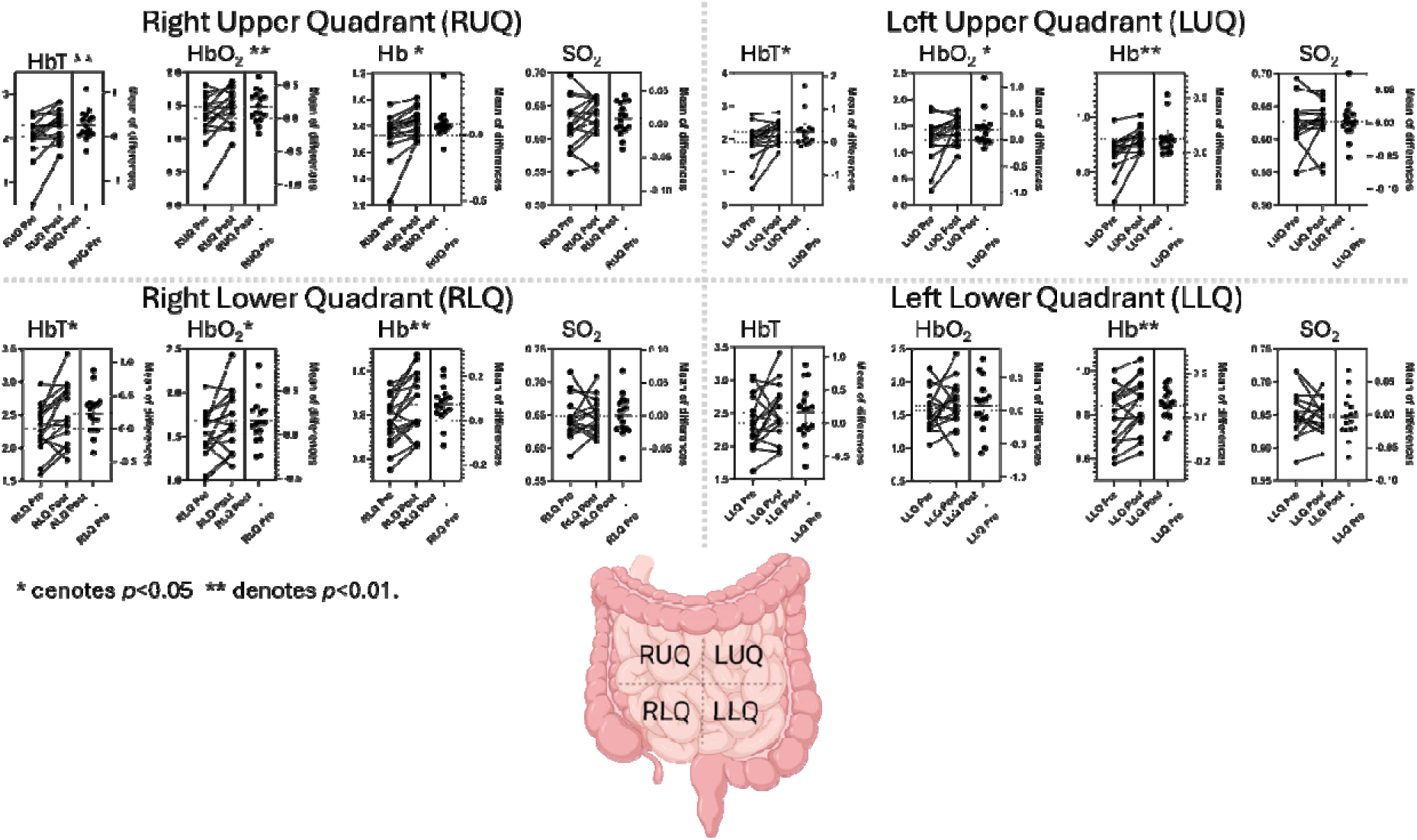
Paired t-tests of pre- and post-prandial measurements for all participants show statistically significant differences in: RUQ, LUQ, and RLQ for total hemoglobin; RUQ, LUQ and RLQ oxyhemoglobin; RUQ, LUQ, RLQ, and LLQ regions for deoxygenated hemoglobin; and none for oxygen saturation. A representation of the abdominal anatomical regions is shown on an abdominal schematic model.

**Figure 6.**
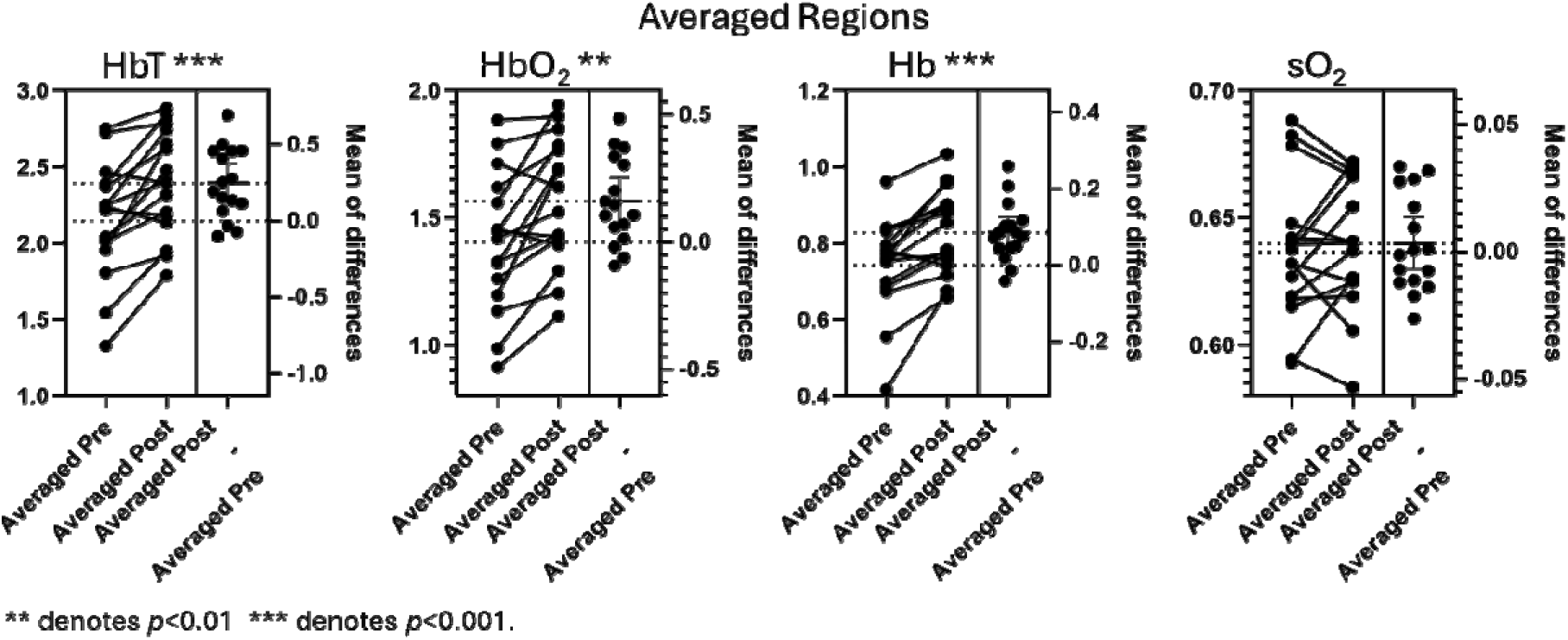
Paired t-tests of pre- and post-prandial measurements for all participants in the region averaged measurement for total hemoglobin, oxyhemoglobin, deoxygenated hemoglobin, and oxygen saturation.

### Secondary Analysis within Age-based Cohorts

Intestinal averages for each PAI measure were clustered into cohorts based on age of the participant: 0-2 years, 3-6 years, and 7-9 years. Quantitative pre- and post-prandial measures for each participant, grouped into age-based cohorts, are shown in Figure 7. In the 7-9-year cohort, with representative images for the cohort in Figure 2, significant values were found for HbT (µ = 0.1632, std ± 0.2775, p < 0.05) and Hb (µ = 0.06973, std ± 0.06295, p < 0.001). In the ages 3-6 cohort, the representative images for the cohort are provided in Figure 3, and significant values were found for HbT (µ = 0.1654, std ± 0.2847, p < 0.01), HbO_2_ (µ = 0.1033, std ± 0.2184, p < 0.05), and Hb (µ = 0.06234, std ± 0.08216, p < 0.01). In the ages 0-2 cohort, representative images for the cohort are provided in Figure 4, and significant values were found for HbT (µ = 0.3642, std ± 0.4757, p < 0.01), HbO_2_ (µ = 0.1033, std ± 0.2184, p < 0.05), and Hb (µ = 0.1089, std ± 0.1433, p < 0.01). No significant values were found for sO_2_ in any cohort.

**Figure 7.**
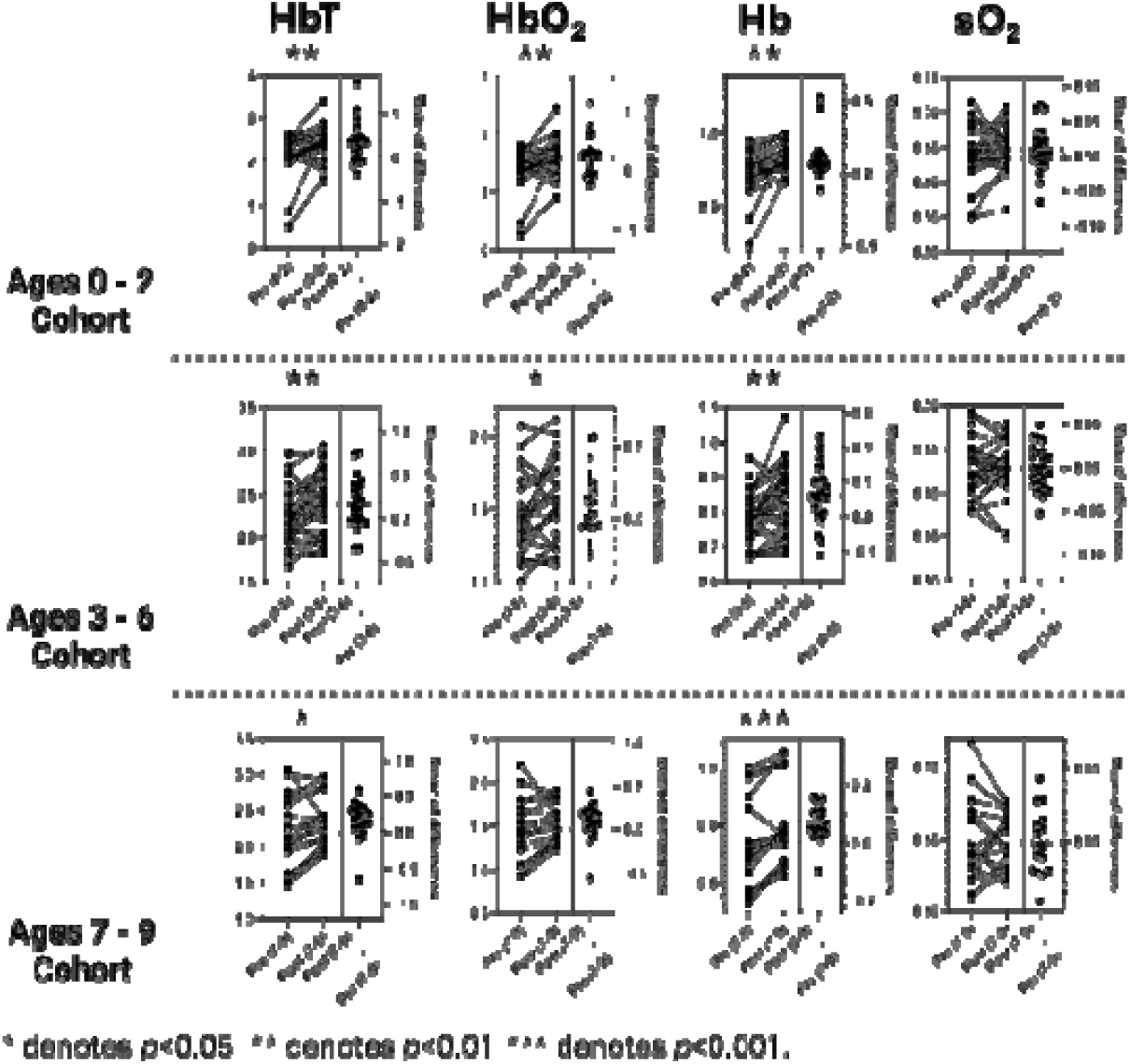
Paired t-tests of pre- and post-prandial measurements of the three cohorts (0­2, 3-6, and 7-9) including all scans. Statistical significance was found for total hemoglobin in the 0-2, 3-6, and 7-9 cohorts, for oxyhemoglobin in the 0-2, and 3-6 cohorts, for deoxygenated hemoglobin in the 0-2, 3-6, and 7-9 cohorts, and none for oxygen saturation.

### Adverse Events

Participants were given a questionnaire for pre-prandial and post-prandial to assess adverse events. Participant responses to questions about AE of the laser safety glasses, imaging probe, and visual assessment results were evaluated. All AE were monitored for symptom occurrence and conclusion. If the symptom did not resolve during the session, there was a follow-up contact to track the event. All AE and resulting statistics are shown in Table 2. No serious adverse events (SAE) occurred during this study. No adverse events relating to the imaging probe were noted. Minor temporary periorbital erythema that resolved within 24 hours was noted in 33% of participants, and 7% noted pressure due to the safety goggles that resolved when removed, with no other adverse safety events reported.

**Table 2.**
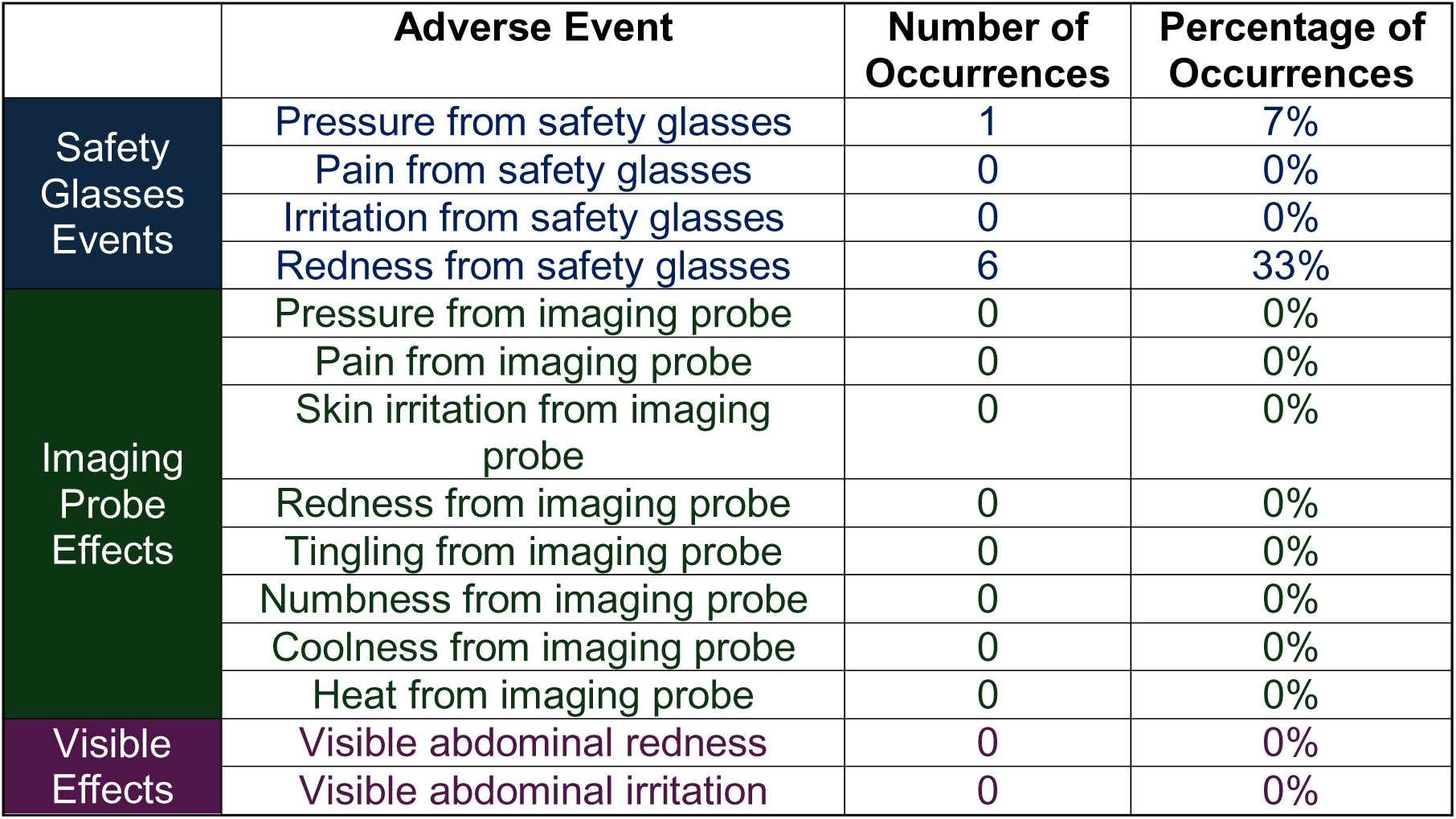
Adverse events that were monitored, number of occurrences, and the total percentage of participants who experienced each adverse event. Most AEs were absent, but 7% experienced pressure and 33% experienced redness from safety glasses. No serious adverse events were reported.

## Discussion

This study demonstrates that abdominal PAI is sensitive to changes in intestinal microvascular circulation following consumption of a lipid-rich snack in pediatric participants. To assess PAI feasibility and safety in pediatric participants, an age ‘step-down’ approach was utilized with cohorts of 7-to-9, 3-to-6, 0-to-2 years of age. We observed significant increases in HbT, Hb, and HbO_2_ between pre- and post-prandial imaging sessions. These changes indicated the expected physiological hyperemic response known to occur in healthy adults and children, as total blood flow increased and increases were non-specific to HbO_2_ or Hb [39]. This validates PAI sensitivity to detect blood flow changes in young pediatric participants. The lack of significant sO_2_ changes may indicate that tissue oxygenation is maintained with feeding in healthy pediatric populations. Combined, these changes are indicative of PAI sensitivity to measure post-prandial hyperemia in various pediatric age groups [38–41]. When examining the safety of abdominal PAI in pediatric participants, we noted no SAEs, and only minor, temporary, and reversible AEs from the laser safety goggles. Our results demonstrate that PAI is safe for pediatric participants.

While our results are promising, there were some minor limitations in this study. Due to the nature of this pilot study, our cohort was limited to healthy pediatric participants. There is a need for future work to assess the clinical NEC population using PAI as an additional non-invasive biomarker to aid in current diagnostic efforts. Our study cohort was also limited in demographic diversity, with limited variation in race. It will be important in future studies to assess PAI in participants with more variation in skin tone, as PAI measures of blood oxygenation can be altered in participants with higher skin melanin concentrations [42, 43]. This concern is notable when considering our ultimate disease application area, as there is a higher incidence of NEC in black patients [42, 43]. Additionally, due to the age step-down approach of our safety and feasibility study design, our study population was limited in age range to a small number of infant participants, which is less representative of the ultimate NICU population. It will be important in future studies to further investigate safety and feasibility in the critical VLBW infant population. Further, the size of the imaging probe also presented a challenge, with probe geometry of approximately 145 x 117 x 51mm, which is relatively large in comparison to infant abdomens, leaving a need for future probe development. Lastly, PAI as an imaging modality is inherently limited by optical penetration depth. In this study, participants over 10 years old were excluded due to low PA signal intensity from body habitus. However, penetration depth was not an issue in participants under 10 years old in our study cohort. As our ultimate patient population for NEC diagnosis is within the VLBW infants, it is important to consider abdominal edema, which is reported to be an increase under 3 mm; since this is within our visualization parameters, we do not anticipate issues [42–44].

PAI is both safe and feasible for assessing abdominal intestinal microvascular function and detecting post-prandial intestinal hyperemia in pediatric participants. Future studies will expand on this pilot study into the clinical NICU patient population to ensure that safety and feasibility are maintained in VLBW premature infants, and to investigate PAI measures of intestinal tissue microvasculature as a diagnostic feature of NEC disease. PAI hardware continues to advance with ongoing technological development and implementation beginning to find clinical applicability and FDA approval [45, 46]. Recent advancements include replacement of solid-state lasers with light-emitting diode (LED) and laser diode-based optical excitation that enhances portability and safety while maintaining suitable penetration depth [47]. As we continue to work past these hurdles, we advance toward the significant promise PAI holds as an emerging noninvasive imaging modality for assessment of intestinal health and disease in pediatric populations.

## Acknowledgements

This work was supported in part by the Wake Forest University School of Medicine CTSI Translational Pilot Program, National Institutes of Health (NIH) R01DK135955, K01DK125633, CTSI UM1TR004929, and AGA Research Scholar Award in Health Disparities. We acknowledge the support of the Wake Forest University School of Medicine CTSI Study Coordinator Pool and Wake Forest University School of Medicine Translational Imaging Program for their support in clinical data collection. We also thank the young participants and parents/guardians for their time and willingness to take part in this research.

## Data availability

The datasets generated and/or analyzed within the current study are available from the corresponding authors on reasonable request.

## Contributions

Study conception and design: JW, VW; Data acquisition: PW, LP, SN, VW, JW; Data analysis and interpretation: PW, LP, VW, JW; Writing and editing the manuscript: All authors.

